# Monoclonal Antibodies for Treatment of SARS-CoV-2 Infection During Pregnancy

**DOI:** 10.1101/2022.04.20.22274090

**Authors:** Erin K. McCreary, Lara Lemon, Christina Megli, Amber Oakes, Rachel L. Zapf, Paula L. Kip, Christopher W. Seymour, the UPMC Magee Monoclonal Antibody Treatment Group

## Abstract

**IMPORTANCE:** Monoclonal antibody (mAb) treatment decreases hospitalization and death in high-risk outpatients with mild to moderate COVID-19. However, no studies have evaluated adverse events and effectiveness of mAbs in pregnant persons compared to no mAb treatment.

**OBJECTIVE:** To determine the frequency of drug-related adverse events and obstetric-associated safety outcomes after treatment with mAb compared to no mAb treatment, and the association between mAb treatment and a composite of 28-day COVID-19-related hospital admission or emergency department visit, COVID-19-associated delivery, or mortality.

**DESIGN, SETTING, PARTICIPANTS:** Propensity-score matched cohort study of persons aged 12 years of age or older with a pregnancy episode and any documented positive SARS-CoV-2 test (polymerase chain reaction or antigen test) in the UPMC health system from April 30, 2021 to January 21, 2022.

**EXPOSURES:** Bamalanivmab and etesevimab, casirivimab and imdevimab, or sotrovimab treatment compared to no mAb treatment.

**MAIN OUTCOMES AND MEASURES:** Drug-related adverse events, obstetric-associated safety outcomes among persons who delivered, and a risk-adjusted composite of 28-day COVID-19-related hospital admission or ED visit, COVID-19-associated delivery, or mortality.

**RESULTS:** Among 944 pregnant persons (median [IQR] age 30 [26, 33] years, White (79.5%, N=750), median [IQR] Charlson Comorbidity Index Score 0 (0,0)), 552 persons received mAb treatment (58%). Median gestational age at COVID-19 diagnosis or treatment was 179 days (IQR: 123, 227), and most persons received sotrovimab (69%, N=382). Of those with known vaccination status, 178 (62%) were fully vaccinated. Drug-related adverse events were uncommon (N=8, 1.4%), and there were no differences in any obstetric-associated outcome among 276 persons who delivered. After propensity score matching, the frequency of the composite 28-day COVID-19-associated outcome was 4.0 per 100 persons (95% CI 1.9, 6.2) in mAb-treated compared to 3.7 per 100 persons (95% CI 1.7, 5.8) in non-treated controls (risk difference = 0.31 per 100 persons [95% CI -2.6, 3.3). There were no deaths among mAb-treated patients compared to 1 death in the non-treated controls (p = 0.24). There were more non-COVID-19-related hospital admissions in the mAb-treated persons (risk difference 2.8 per 100 persons (95% CI 1.1, 4.5)).

**CONCLUSIONS AND RELEVANCE:** In pregnant persons with mild to moderate COVID-19, adverse events after mAb treatment were mild and rare. There was no difference in obstetric-associated safety outcomes between mAb treatment and no treatment among persons who delivered. MAb treatment was associated with similar 28-day COVID-19-associated outcomes and more non-COVID-19-related hospital admissions compared to no mAb treatment.

**Key Points:** *QUESTION:* Among pregnant persons with COVID-19, is monoclonal antibody (mAb) treatment associated with drug-related adverse events, similar frequency of obstetric-associated safety outcomes, and improved COVID-19-related clinical outcomes compared to no mAb treatment?

*FINDINGS:* In 944 pregnant persons with COVID-19, drug-related adverse events were mild and infrequent. Obstetric-associated safety outcomes were similar between mAb treatment and no treatment. There was no evidence of difference in risk of COVID-19-related hospital admission, COVID-19-associated delivery, or mortality between mAb treatment and no mAb treatment.

*MEANING:* In pregnant persons with mild to moderate COVID-19, adverse events after mAb treatment were uncommon, and there was no difference in obstetric-associated safety outcomes between mAb treatment and no treatment. MAb treatment was associated with similar 28-day risk of a COVID-19-associated outcome and more non-COVID-19-related hospital admissions compared to no mAb treatment.

## Background

Monoclonal antibody (mAb) treatment is associated with decreased hospitalization and death in outpatients with mild to moderate COVID-19.^1-3^ The Emergency Use Authorizations (EUA) for these compounds were updated in May 2021 to list pregnancy as a risk factor for progression to severe disease.^3-6^ Yet, no studies evaluate the effectiveness of mAbs targeting SARS-CoV-2 among pregnant persons, data are limited on the safety of mAbs in pregnancy, and immunoglobulin-based therapy remains controversial.^7-13^

The purpose of this study is to estimate the rate of drug-related adverse events from mAb treatment among a cohort of pregnant persons with SARS-CoV-2 infection, the rate of obstetric-associated safety outcomes among all persons who delivered, and the risk-adjusted association between mAb treatment and composite 28-day COVID-19-related hospital admission or emergency department (ED) visit, COVID-19-associated delivery, or mortality compared to no mAb treatment.

## Methods

This study was approved by the UPMC Quality Improvement Review Committee and University of Pittsburgh Institutional Review Board. Methods and results are reported in accordance with the Reporting of Studies Conducted Using Observational Routinely Collected Health Data (RECORD) statement.^14^ The study followed the Strengthening the Reporting of Observational Studies in Epidemiology (STROBE) reporting guideline **(Supplement 2)**.^15^

This was a cohort study of mAb EUA-eligible persons (aged 12 years of age or older) with a pregnancy episode and any documented positive SARS-CoV-2 test (polymerase chain reaction or antigen test) in the UPMC health system from April 30, 2021 to January 21, 2022.

Prior to December 23, 2021, all mAb-treated patients received mAb via a central management and allocation system which has been previously described.^16-19^ A small minority of patients received subcutaneous casirivimab and imdevimab to accommodate surging patient referrals and staffing shortages.^20^ From December 23, 2021 through January 21, 2022, all patients received intravenous sotrovimab due to the emergence of the Omicron variant. Starting September 28, 2021, pregnant patients and patients with immunocompromised conditions were given priority for mAb treatment due to drug scarcity. Patients reviewed the US Food and Drug Administration EUA Fact Sheet(s) and verbally consented to treatment with any available mAb prior to mAb administration.

Patients were considered mAb-treated if they received any mAb in an outpatient infusion center, urgent care facility, or obstetric triage area and non-treated if they did not receive mAb. Patients were excluded if they had no previous care at UPMC in the preceding 12 months, received mAb for post-exposure prophylaxis, or received mAb while inpatient status or in a non-obstetric ED. The obstetric ED was used as an outpatient referral center during this time period and therefore the persons treated there were included in this analysis as their clinical status was deemed ‘outpatient’ rather than ‘emergency department’. To account for immortal time bias between SARS-CoV-2 testing and mAb treatment, non-treated persons with a hospital admission within 1 day of their positive SARS-CoV-2 test result were excluded.^21^ Both groups required 28-day follow-up. For non-treated control subjects, the 28-day outcome ascertainment period started on the day after the SARS-CoV-2 test positive result. For treated subjects, the 28-day outcome ascertainment period started on the day of mAb treatment.

### Outcomes

The primary descriptive safety outcomes were the rates of drug-related adverse events reported by patients or providers at each treatment site among persons who received mAb and obstetric-associated outcomes [i.e., gestational age at delivery, birthweight, stillbirth, neonatal intensive care unit (NICU) admission, diagnosis of hypertension at time of delivery, severe maternal morbidity (defined in accordance with CDC guidelines), and maternal ICU visit] for all persons that delivered in the study time-frame.^22^ To determine mAb effectiveness, the primary outcome was the risk-adjusted association between mAb treatment and a composite of 28-day COVID-19-related hospital admission or ED visit, COVID-19-associated delivery, or mortality. A COVID-19-related hospital admission was defined as an antepartum admission for supportive oxygen or additional respiratory support. A COVID-19-associated delivery was identified as a delivery indicated secondary to COVID-19-related complications including, i.) maternal respiratory failure, or ii.) fetal distress with evidence of pathognomonic SARS-CoV-2-placentitis.^23-25^ Secondary effectiveness outcomes included 28-day non-COVID-19 related admissions and rates of individual components of the composite outcome.

### Data Sources and Definitions

Demographic, obstetric, and clinical data were abstracted from the UPMC Clinical Data Warehouse (CDW). The CDW stores all discrete data entered into each electronic medical record across the health system. Race was patient-identified and classified as Black, White, or Other (included Alaska Native, American Indian, Asian, Filipino, Indian, Native Hawaiian, or Pacific Islander). Dates entered in pregnancy episodes were used to calculate the estimated gestational age at the time of positive SARS-CoV-2 test or mAb treatment. Sociodemographic data, medical history, and administrative claims data for all outpatient and in-hospital encounters were collected with diagnoses and procedures coded based on the International Classification of Diseases, Ninth and Tenth revisions (ICD-9 and ICD-10, respectively).^26,27^

Adverse events were defined as any reaction that occurred during the observation period after mAb injection or infusion (e.g., rash, shortness of breath, etc.) and were recorded by practitioners at each infusion center in a secure electronic file-sharing application. Nursing and physician staff also used an internal, nonpunitive, patient safety reporting system (“Risk Master”) for adverse reactions and medication errors. Delivery outcomes were recorded in Delivery Forms in the medical record.

The primary effectiveness outcome was identified using hospitalizations and ED visits from the CDW. Hospital discharge disposition of “Ceased to Breathe” corresponded to in-hospital mortality, and deaths after discharge were identified with the Death Master File from the Social Security Administration.^28^ Detailed chart review was performed by a subspecialist in Maternal-Fetal Medicine (C.M.) to stratify admission and delivery indication for the primary and secondary outcomes. The study team (C.M.) reviewed cases with estimated gestational age <98 days at the time of event to identify early miscarriages. Vaccination status was confirmed by a clinical pharmacist (A.O.) using the electronic medical record, medical referral orders, and Pennsylvania Statewide Immunization Information System (PA-SIIS).

### Statistical Analyses

Baseline characteristics and adverse events were compared by treatment status using chi-2 testing, Fisher’s Exact tests, unpaired t-tests, and Wilcoxon Rank-Sum testing, as appropriate. To understand the risk-adjusted association between treatment and outcome, a multivariable logistic regression with three analytic approaches was used, i.) conditionally in a propensity score-matched sample, ii.) conditionally in an inverse probability weighted sample based on the propensity score, and iii.) adjusting for the propensity score. Results are presented as predicted risk per 100 pregnant people, the risk difference between the treated and non-treated, and as adjusted risk ratios with 95% confidence intervals.

Propensity scores were generated using multivariate logistic regression modeling the likelihood of receiving mAb treatment. Variables included in the model were decided *a priori* and based on clinical judgement, including age, race, estimated gestational age, and vaccination status at time of event, month and year of event, insurance type (commercial, public, self-pay), area deprivation index score, parity, gravidity, diagnosis of preexisting hypertension, preeclampsia, chronic hypertension, pre-gestational diabetes, gestational diabetes, asthma, cancer, rheumatoid arthritis, infertility, anxiety, depression, atrial fibrillation, chronic heart failure, irritable bowel syndrome, hyperlipidemia, use of corticosteroids, history of chemotherapy, tobacco use, alcohol use, body mass index, and blood pressures at the closest office visit within one year prior, and indicators for missingness of any data. The timing of the positive SARS-CoV-2 result or treatment date was included in the propensity score model to account for unmeasured differences in variant waves. Missing data were replaced with the respective group’s median value, and indicators of missingness were included in the propensity score.

Propensity score distributions were assessed to ensure balance across treatment groups. Propensity score matching in a 1:1 ratio was performed with common support and nearest-neighbor in a caliper of 0.03. Covariates were compared before and after matching to ensure standardized differences of less than 10%. In the propensity score-matched cohort, logit models and adjusted predictive margins were used. In the population with common support and inverse probability weights, logit models accounted for weighting to model the primary outcome and margins to determine the adjusted predicted risk and risk difference. Finally, COVID-associated visit was modeled using logit models adjusting for the propensity score and calculated the associated risk.

### Post hoc exploratory analyses

To understand how treatment may be different in subgroups of pregnant persons, we stratified the population by vaccination status (unvaccinated vs fully vaccinated) and obesity status (obese vs non-obese) at time of event. In these subpopulations we first conducted crude logistic regressions modeling likelihood of 28-days COVID-associated admission, then conditionally adjusting for the propensity scores. This was repeated exploring the secondary outcome of non-COVID admission within 28 days of event. Results are again presented as predicted risk per 100 pregnant people, the risk difference between the treated and non-treated, and as adjusted risk ratios with 95% confidence intervals. All analyses were performed in Stata IC, version 16 software package (StataCorp LLC), and statistical significance corresponded to a p<0.05.

## Results

### Study population

Of 1,140 pregnant persons with a positive SARS-CoV-2 test, 944 (83%) were included in the cohort, among whom 552 (58%) were treated with mAb. Patients were primarily excluded if receiving mAb treatment in a non-OB ED (n=86, 7.5%), inpatient admission (n=44, 3.8%), or were hospitalized within 24 hours of positive test (n=56, 4.9%). Most patients were young (median age 30 years [IQR: 26,33]), White (79.5%, n=750), and had few comorbidities (median Charlson Comorbidity Index 0 [IQR: 0,0]). Of those with known vaccination status, 178 (62%) were fully vaccinated. MAb-treated patients were older (median age 30.0 vs 28.9 years, P <0.001), more likely to have commercial insurance (67.9% vs 56.4%, P <0.001), and had a lower median area of deprivation index score (64 [IQR: 44,82] vs 72 [IQR: 54,88], P <0.001). MAb-treated patients were more likely to have a history of infertility (12.3% vs 6.6%, P = 0.004) and to be fully vaccinated (48.0% vs 32.4%, p<0.001). The median gestational age at time of COVID-19 diagnosis or treatment was 179 days (IQR: 122.5, 226.5). Among treated persons, most (58%, n=320) received mAb within 4 days of symptom onset. The most common mAb was sotrovimab (69%, n=382) compared to casirivimab and imdevimab (20%, n=110) and bamlanivimab and etesevimab (11%, n=60). The mean (SD) time from SARS-CoV-2 test result to mAb treatment was 1 (±6) day **(Table 1)**.

**Table 1.**
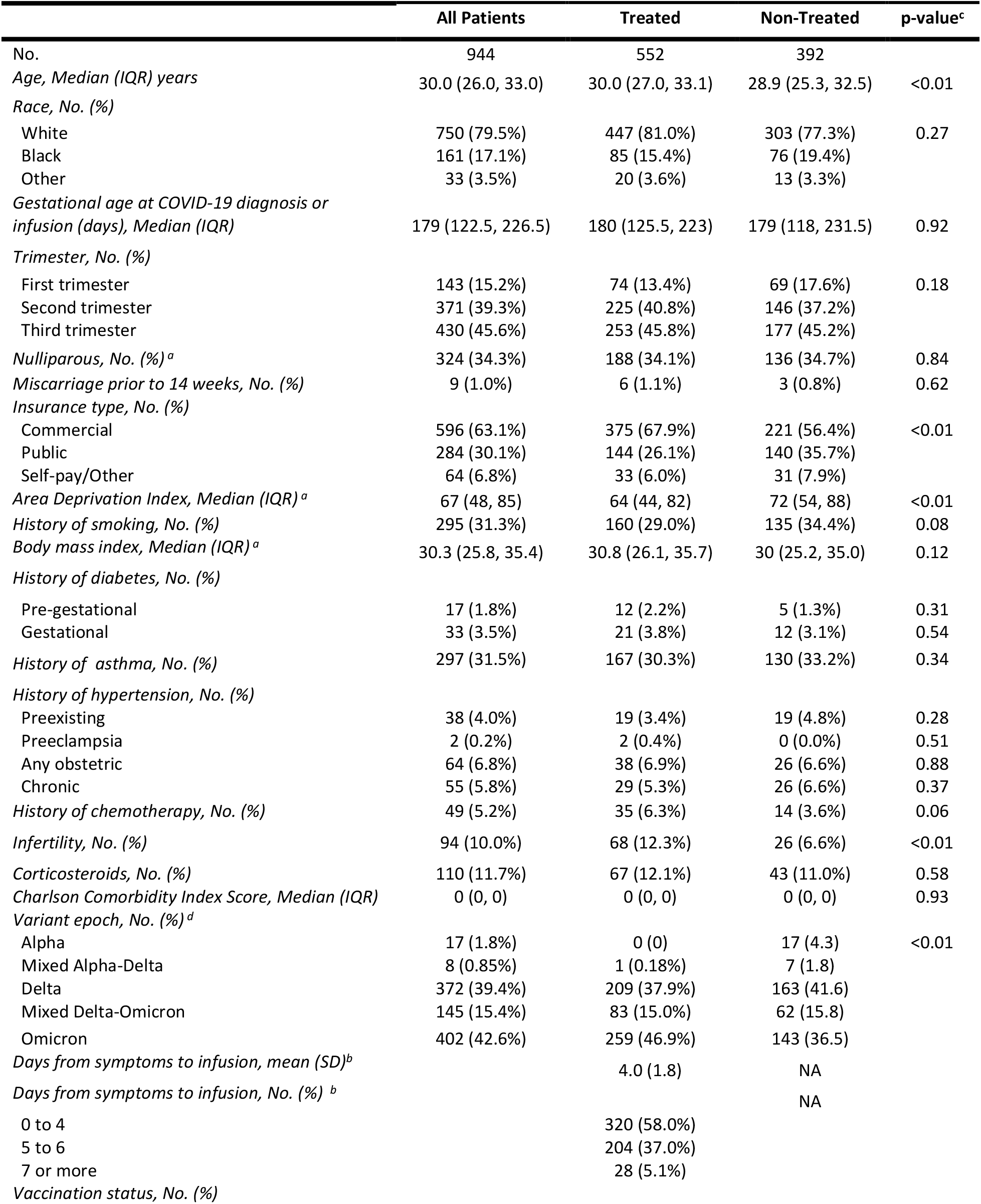
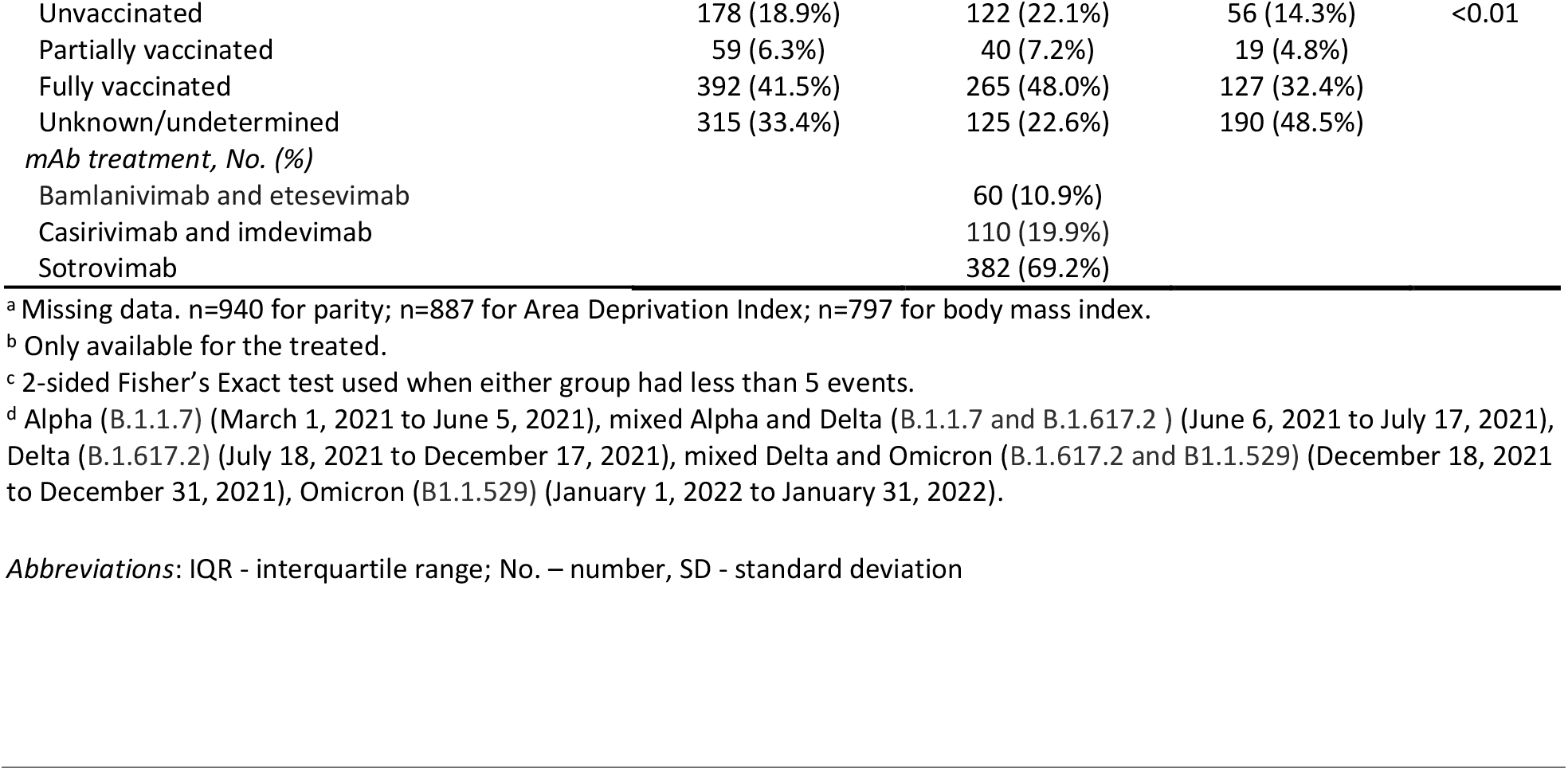
Comparison of Characteristics of Pregnant mAb Treated vs. Non-treated Persons.

|  | All Patients | Treated | Non-Treated | p-value<sup>c</sup> |
| --- | --- | --- | --- | --- |
| No. | 944 | 552 | 392 |  |
| Age, Median (IQR) years | 30.0 (26.0, 33.0) | 30.0 (27.0, 33.1) | 28.9 (25.3, 32.5) | <0.01 |
| Race, No. (%) |  |  |  |  |
| White | 750 (79.5%) | 447 (81.0%) | 303 (77.3%) | 0.27 |
| Black | 161 (17.1%) | 85 (15.4%) | 76 (19.4%) |  |
| Other | 33 (3.5%) | 20 (3.6%) | 13 (3.3%) |  |
| Gestational age at COVID-19 diagnosis or infusion (days), Median (IQR) | 179 (122.5, 226.5) | 180 (125.5, 223) | 179 (118, 231.5) | 0.92 |
| Trimester, No. (%) |  |  |  |  |
| First trimester | 143 (15.2%) | 74 (13.4%) | 69 (17.6%) | 0.18 |
| Second trimester | 371 (39.3%) | 225 (40.8%) | 146 (37.2%) |  |
| Third trimester | 430 (45.6%) | 253 (45.8%) | 177 (45.2%) |  |
| Nulliparous, No. (%)<sup>a</sup> | 324 (34.3%) | 188 (34.1%) | 136 (34.7%) | 0.84 |
| Miscarriage prior to 14 weeks, No. (%) | 9 (1.0%) | 6 (1.1%) | 3 (0.8%) | 0.62 |
| Insurance type, No. (%) |  |  |  |  |
| Commercial | 596 (63.1%) | 375 (67.9%) | 221 (56.4%) | <0.01 |
| Public | 284 (30.1%) | 144 (26.1%) | 140 (35.7%) |  |
| Self-pay/Other | 64 (6.8%) | 33 (6.0%) | 31 (7.9%) |  |
| Area Deprivation Index, Median (IQR)<sup>a</sup> | 67 (48, 85) | 64 (44, 82) | 72 (54, 88) | <0.01 |
| History of smoking, No. (%) | 295 (31.3%) | 160 (29.0%) | 135 (34.4%) | 0.08 |
| Body mass index, Median (IQR)<sup>a</sup> | 30.3 (25.8, 35.4) | 30.8 (26.1, 35.7) | 30 (25.2, 35.0) | 0.12 |
| History of diabetes, No. (%) |  |  |  |  |
| Pre-gestational | 17 (1.8%) | 12 (2.2%) | 5 (1.3%) | 0.31 |
| Gestational | 33 (3.5%) | 21 (3.8%) | 12 (3.1%) | 0.54 |
| History of asthma, No. (%) | 297 (31.5%) | 167 (30.3%) | 130 (33.2%) | 0.34 |
| History of hypertension, No. (%) |  |  |  |  |
| Preexisting | 38 (4.0%) | 19 (3.4%) | 19 (4.8%) | 0.28 |
| Preeclampsia | 2 (0.2%) | 2 (0.4%) | 0 (0.0%) | 0.51 |
| Any obstetric | 64 (6.8%) | 38 (6.9%) | 26 (6.6%) | 0.88 |
| Chronic | 55 (5.8%) | 29 (5.3%) | 26 (6.6%) | 0.37 |
| History of chemotherapy, No. (%) | 49 (5.2%) | 35 (6.3%) | 14 (3.6%) | 0.06 |
| Infertility, No. (%) | 94 (10.0%) | 68 (12.3%) | 26 (6.6%) | <0.01 |
| Corticosteroids, No. (%) | 110 (11.7%) | 67 (12.1%) | 43 (11.0%) | 0.58 |
| Charlson Comorbidity Index Score, Median (IQR) | 0 (0, 0) | 0 (0, 0) | 0 (0, 0) | 0.93 |
| Variant epoch, No. (%)<sup>d</sup> |  |  |  |  |
| Alpha | 17 (1.8%) | 0 (0) | 17 (4.3) | <0.01 |
| Mixed Alpha-Delta | 8 (0.85%) | 1 (0.18%) | 7 (1.8) |  |
| Delta | 372 (39.4%) | 209 (37.9%) | 163 (41.6) |  |
| Mixed Delta-Omicron | 145 (15.4%) | 83 (15.0%) | 62 (15.8) |  |
| Omicron | 402 (42.6%) | 259 (46.9%) | 143 (36.5) |  |
| Days from symptoms to infusion, mean (SD)<sup>b</sup> |  | 4.0 (1.8) | NA |  |
| Days from symptoms to infusion, No. (%)<sup>b</sup> |  |  | NA |  |
| 0 to 4 |  | 320 (58.0%) |  |  |
| 5 to 6 |  | 204 (37.0%) |  |  |
| 7 or more |  | 28 (5.1%) |  |  |
| Vaccination status, No. (%) |  |  |  |  |
| Unvaccinated | 178 (18.9%) | 122 (22.1%) | 56 (14.3%) | <0.01 |
| Partially vaccinated | 59 (6.3%) | 40 (7.2%) | 19 (4.8%) |  |
| Fully vaccinated | 392 (41.5%) | 265 (48.0%) | 127 (32.4%) |  |
| Unknown/undetermined | 315 (33.4%) | 125 (22.6%) | 190 (48.5%) |  |
| *mAb treatment, No. (%)* |  |  |  |  |
| Bamlanivimab and etesevimab |  | 60 (10.9%) |  |  |
| Casirivimab and imdevimab |  | 110 (19.9%) |  |  |
| Sotrovimab |  | 382 (69.2%) |  |  |

<sup>a</sup> Missing data. n=940 for parity; n=887 for Area Deprivation Index; n=797 for body mass index.
<sup>b</sup> Only available for the treated.
<sup>c</sup> 2-sided Fisher's Exact test used when either group had less than 5 events.
<sup>d</sup> Alpha (B.1.1.7) (March 1, 2021 to June 5, 2021), mixed Alpha and Delta (B.1.1.7 and B.1.617.2) (June 6, 2021 to July 17, 2021), Delta (B.1.617.2) (July 18, 2021 to December 17, 2021), mixed Delta and Omicron (B.1.617.2 and B.1.529) (December 18, 2021 to December 31, 2021), Omicron (B.1.529) (January 1, 2022 to January 31, 2022).
*Abbreviations:* IQR - interquartile range; No. – number, SD - standard deviation

### Adverse events from mAb treatment and obstetric-associated safety outcomes

Drug-related adverse events were mild and occurred in 8 patients treated with mAb (1.4%). No patients experienced a severe infusion-related reaction. Among the 276 persons who delivered in the follow-up period, there were no differences between groups for gestational age at delivery, birthweight, neonatal intensive care unit admission, stillbirths, severe maternal morbidity, hypertension at delivery, or maternal ICU admission **(Table 2, Table 3)**.

**Table 2.**
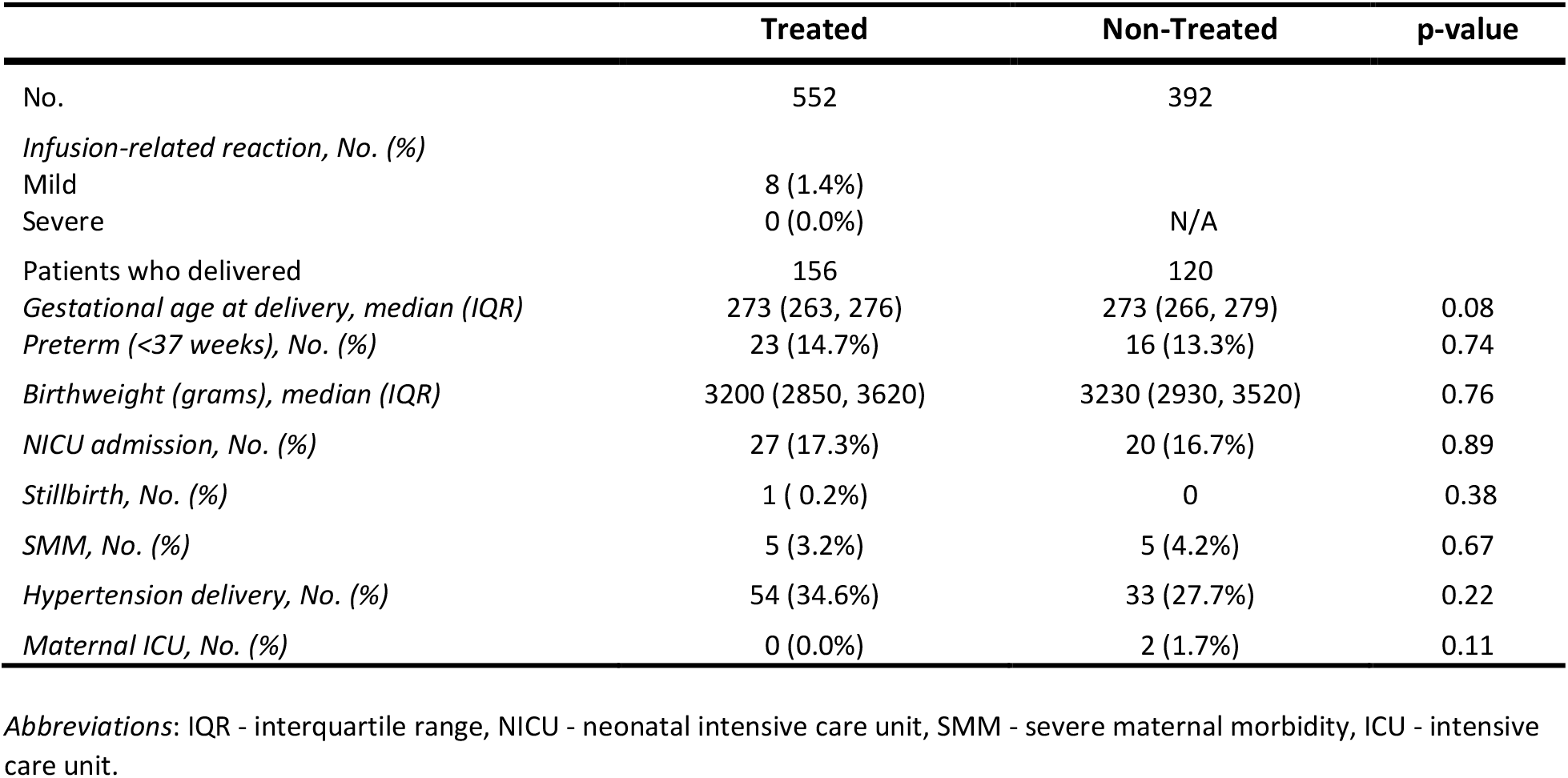
Safety of Monoclonal Antibody Treatment in Pregnant Persons.

**Table 3.** Detailed outcomes within 28 days of test or treatment in Pregnant mAb Treated vs. Non-treated Patients.

|  | All Patients | Treated | Non-Treated | p-value<sup>a</sup> |
| --- | --- | --- | --- | --- |
| No. | 944 | 552 | 392 |  |
| *Composite outcome, No. (%)* | 34 (3.6%) | 17 (3.1%) | 17 (4.3%) | 0.31 |
| *COVID-associated admission, No. (%)* | 4 (0.4%) | 2 (0.4%) | 2 (0.5%) | 1.0 |
| *COVID-associated delivery, No (%)* | 4 (0.4%) | 1 (0.2%) | 3 (0.8%) | 0.31 |
| *COVID-associated ED/Triage visit, No (%)* | 29 (3.1%) | 14 (2.5%) | 15 (3.8%) | 0.26 |
| *Death, No. (%)* | 1 (0.1%) | 0 (0%) | 1 (0.3%) | 0.42 |
| *Other admission, No. (%)* | 16 (1.7%) | 14 (2.5%) | 2 (0.5%) | 0.017 |
| Hypertension, No. (%) | 1 (0.1%) | 1 (0.2%) | 0 (0%) |  |
| Hyperemesis, No. (%) | 1 (0.1%) | 1 (0.2%) | 0 (0%) |  |
| Preterm contractions, No. (%) | 2 (0.2%) | 2 (0.4%) | 0 (0%) |  |
| Pyelonephritis, No. (%) | 3 (0.3%) | 2 (0.4%) | 1 (0.3%) |  |
| Nephrolithiasis, No. (%) | 1 (0.1%) | 0 (0%) | 1 (0.3%) |  |
| Intrahepatic cholestasis of pregnancy, No. (%) | 2 (0.2%) | 2 (0.4%) | 0 (0%) |  |
| Intrauterine growth restriction, No. (%) | 1 (0.1%) | 1 (0.2%) | 0 (0%) |  |
| Seizure, No. (%) | 1 (0.1%) | 1 (0.2%) | 0 (0%) |  |
| Preterm premature rupture of membranes, No. (%) | 1 (0.1%) | 1 (0.2%) | 0 (0%) |  |
| Vaginal bleeding, No. (%) | 1 (0.1%) | 1 (0.2%) | 0 (0%) |  |
| Hyperglycemia, No. (%) | 1 (0.1%) | 1 (0.2%) | 0 (0%) |  |
| Burn, No. (%) | 1 (0.1%) | 1 (0.2%) | 0 (0%) |  |
| *Delivery admission, No. (%)* | 135 (14.3%) | 82 (14.9) | 53 (13.5) | 0.56 |

<sup>a</sup> 2-sided Fisher's Exact test used when either group had less than 5 events.
Abbreviations: No. - number

### Association of mAb treatment with outcome

Among 648 matched patients (324 treated and untreated), clinical characteristics were similar between groups **(eTable 2, eFigure 2 in the supplement)**. Prior to matching, more patients were treated in the later months of the study period, although (i.e., presumed SARS-CoV-2 variant) was similar after matching (**eFigure 3 in the supplement**). In the primary analysis in the matched cohort, the composite 28-day risk-adjusted frequency of a COVID-19-associated outcome was 4.0 per 100 persons (95% CI 1.9, 6.2) in mAb-treated compared 3.7 per 100 persons (95% CI 1.7, 5.8) in non-treated controls (risk difference 0.31 per 100 persons [95% CI -2.6, 3.3]). The propensity score-adjusted risk ratio was 1.0 (95%CI 0.5, 2.0) for mAb treated compared to untreated persons. The results were similar using the inverse probability weighted sample based on the propensity score (risk ratio 1.04 (95%CI 0.52, 2.1) and propensity score-matched models (risk ratio 1.08 (95%CI 0.5, 2.3) (**Table 4**).

**Table 4.**
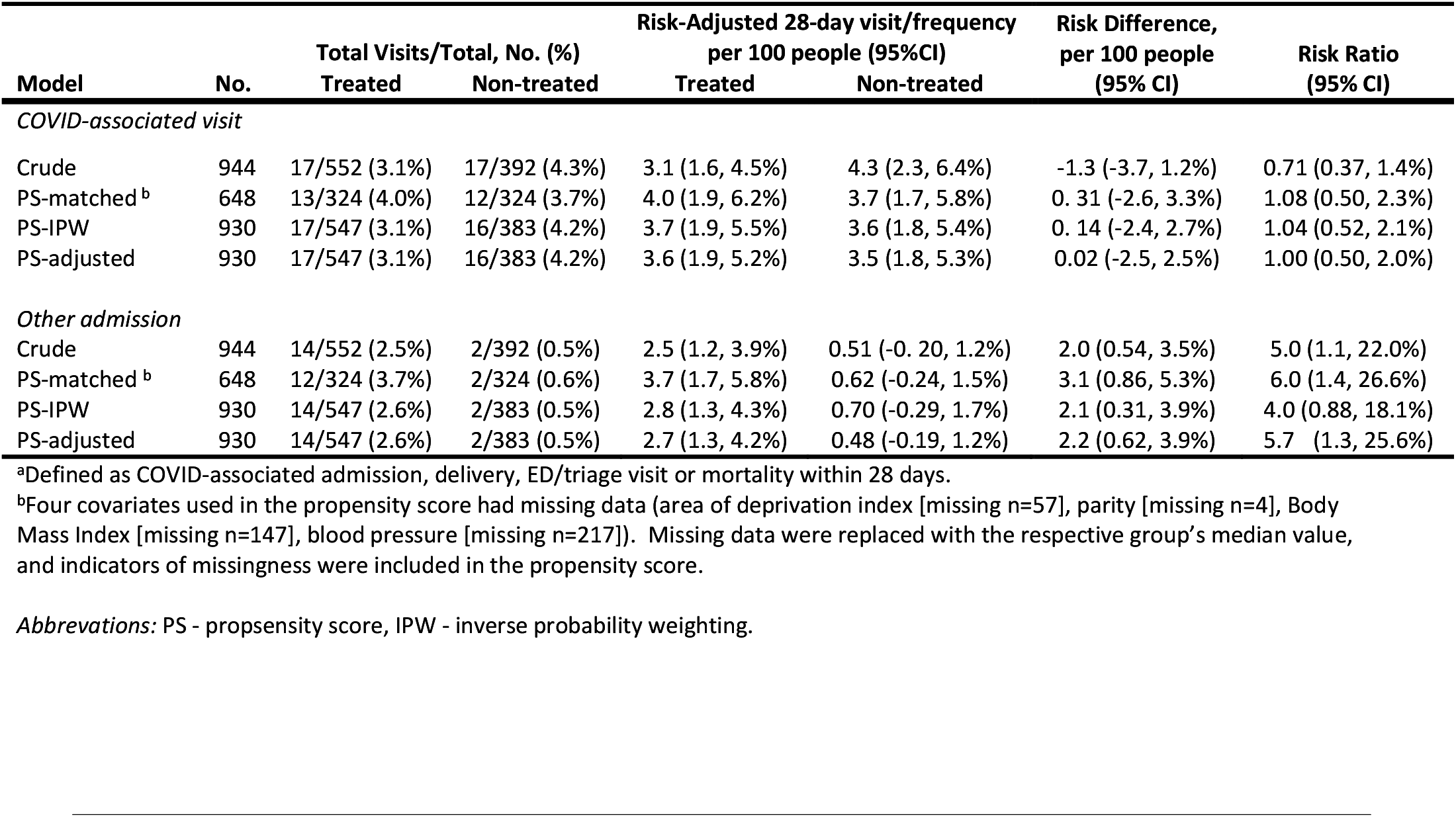
Risk-Adjusted Frequency of COVID-19 Associated Outcome or Other Admission Within 28-days of SARS-CoV-2 positive Test or Monoclonal Antibody Infusion.

The individual components of the composite outcome were similar between groups **(eTable 1 in the supplement)**. There were no deaths among mAb-treated patients compared to 1 death in the non-treated controls (p = 0.24). There were more non-COVID-19-related hospital admissions in the mAb-treated patients [14 (2.5%) vs 2 (0.5%), P = 0.017, and the risk difference in the propensity-adjusted cohort was 2.8 per 100 people (95% CI 1.1, 4.5), **Table 3, Table 4)**. Common indications non-COVID-19 admissions were preterm contractions (N=2, 11%) and intrahepatic cholestasis of pregnancy (N=2, 11%).

In post hoc exploratory analyses among the subset of vaccinated patients (N=392), the composite 28-day propensity-adjusted, risk-adjusted frequency of a COVID-19-associated outcome was 2.6 per 100 persons (95% CI 0.53, 4.7) in mAb-treated compared to 0.63 per 100 persons (95% CI -0.61, 1.9) in non-treated controls (risk difference 2 per 100 persons [95% CI -0.47, 4.4], **eTable 2 and eFigure 1 in the supplement**). For unvaccinated patients, the composite 28-day propensity-adjusted, risk-adjusted frequency of a COVID-19-associated outcome was 4.2 per 100 persons (95% CI 0.61, 7.8) in mAb-treated compared to 10.2 per 100 persons (95% CI 2.5, 18) in non-treated controls (risk difference -6 per 100 persons [95% CI -14.5, 2.5]).

## Discussion

In a cohort study of 944 pregnant persons, drug-related adverse events following mAb treatment were rare and there was no difference in obstetric-associated outcomes among persons who delivered. The risk-adjusted frequency of 28-day COVID-19-associated outcomes for mAb treatment compared to no mAb treatment was similar; however, there were more non-COVID-19-related hospital admissions in mAb-treated patients.

Prior work reported on the use of mAbs in pregnant persons with COVID-19 in small case series. These data demonstrate no serious treatment-related adverse events. However, a non-treated comparator was not reported. We expand on this work with the largest safety and effectiveness evaluation of mAb treatment compared to non-treatment in pregnant persons to our knowledge, including both a description of clinically adjudicated adverse events and COVID-19-related outcomes. Importantly, mAb treatment appears safe in pregnancy with respect to drug-related adverse events and obstetric-associated outcomes (i.e., gestational age at delivery, birthweight, stillbirth, neonatal intensive care unit admission, hypertension, severe maternal morbidity, and maternal intensive care unit visit).

While COVID-19-associated outcomes were similar between groups, the event frequency for hospitalization and death were lower than previously reported frequencies for the general population for both mAb-treated and non-treated groups. The low event frequency may reflect the younger population with few comorbidities and inclusion of fully vaccinated patients in the cohort. The neutral finding from effectiveness models may be due to the sample size of the cohort or the absence of a true treatment effect. Our study is unable to evaluate whether mAbs are associated with a difference in risk-adjusted COVID-19-related outcomes for pregnant persons at higher risk of COVID-19-related complications, such as those with multiple comorbidities, other immunocompromising conditions, and/or unvaccinated status.

MAb-treated patients experienced significantly more non-COVID-19 admissions compared to patients who did not receive mAb treatment. We are unable to determine if these admissions were related to mAb treatment or not. A time-to-event analysis revealed only 2 admissions within the first week of receiving mAb, one seizure in a patient with known epilepsy and 1 episode of hyperglycemia **(eFigure 1 in the supplement)**. The absolute difference in non-COVID hospitalizations (14 events versus 2 events) suggests unmeasured differences may be present between groups unrelated to mAb treatment. The mAb treated patients had higher rates of vaccination, access to fertility care, and more commercial insurance, and therefore may be more likely to access health care for a non-COVID-19 reason. Monitoring bias, when providers followed patients who received mAb more closely than those who did not receive mAb, may also have contributed to the finding. Importantly, this study did not show a difference in hypertension-related, preterm deliveries, or severe maternal morbidity.

Recent clinical practice guidelines from the National Institutes of Health, Centers for Disease Control and Prevention, American College of Obstetricians and Gynecologists, and Society for Maternal-Fetal Medicine recognize pregnant persons as higher risk for severe disease from SARS-CoV-2 infection.

Severe disease is most common in pregnant people aged 35 to 44 years compared to younger patients, and COVID-19 can increase risk of preterm birth or stillbirth amongst all pregnant people. However, these data derive from the Alpha and Delta variant eras. More recent data suggest that those with mild to moderate disease do not have increased rates of adverse neonatal outcomes, and that Omicron-infected persons experience less severe disease than persons infected with previous variants.^29,30^ In the meantime, current guidelines recommend *against* withholding treatment for COVID-19, including mAb, from pregnant or lactating individuals because of theoretical safety concerns.^29^ In the context of this study, routine use of mAbs in pregnant persons with minimal comorbidities and low risk of severe disease in the Omicron variant era may not benefit from treatment. However, it is unknown if mAbs would benefit (or harm) pregnant persons with additional risk factors for severe disease, and if different mAbs are variably effective against different SARS-CoV-2 variants in pregnant persons.

The study has several limitations. First, drug-related adverse events were patient and provider reported and potentially underrepresented. Second, a minority of patients completed pregnancy within the follow up period, limiting data on delivery complications and neonatal outcomes. Third, symptom severity at the time of testing and treatment (whether symptomatic or asymptomatic) was not available for non-treated patients. However, the average time from SARS-CoV-2 test result to treatment was 1 day, suggesting immortal time bias between mAb-treated and non-treated is very unlikely. Fourth, as with any observational study, these findings do not provide causal inference, as many unmeasured confounders may be present. We used multiple modeling approaches and found consistent results.

Fifth, most patients in the cohort received sotrovimab when the Omicron variant was dominant in our geographic region. These data may not be generalizable to other variants, regions, or time periods.

## Conclusion

In pregnant persons with mild to moderate COVID-19, adverse events after mAb treatment were mild and rare. There was no difference in obstetric-associated safety outcomes between mAb treatment and no mAb treatment among persons who delivered. MAb treatment was associated with similar 28-day COVID-19-associated outcomes and more non-COVID-19-related hospital admissions compared to no mAb treatment.

## Data Availability

All data produced in the present study are available upon reasonable request to the authors.

## Article Information

## Acknowledgements

We acknowledge staff at UPMC Clinical Analytics, the UPMC Wolff Center, and Biostatistics and Data Management Core at the CRISMA Center in the Department of Critical Care Medicine at the University of Pittsburgh for curating and managing the data. The authors also thank the clinical staff of the UPMC monoclonal antibody infusion centers as well as the support and administrative staff behind this effort, including but not limited to: Jodi Ayers, Roshni Bag, Ashley Beyerl, Trudy Bloomquist, Patty Boland, Mikaela Bortot, Jonya Brooks, Julie Brown, James Cable, Sherry Casali, Donna Cochran, Kate Codd-Palmer, Jeana Colella, Amy Cooper, Jennifer Dueweke, Jesse Duff, Janice Dunsavage, Jessica Fesz, Kathleen Flinn, Daniel Gessel, Jennifer Gutshall, Shawn Gronlund, Amy Helmuth, Erik Hernandez, Larry Hruska, Rosella Hoffman, Allison Hydzik, Le Ann Kaltenbaugh, LuAnn King, Jim Krosse, Sheila Kruman, Zachary Lenhart, Amy Lukanski, Sharon Mackall, Hilary Maskiewicz, Debra Masser, Rebecca Medva, Jenny Lynne Morris, Theresa Murillo, Christine O’Neill, Carolyn Persichetti, Lea Plagens, Melanie Pierce, Teressa Polcha, Kevin Pruznak, Dejeana Rawlins, Debra Rogers, Johnanne Ross, Rozalyn Russell, Samantha Sacco, Sarah Sakaluk, Heather Schaeffer, Robert Shulik, Libby Shumaker, Susan Spencer, Betsy Tedesco, Troy Treanor, Ken Trimmer, Brehan Wolff, Shannon Work, Jennifer Zabala, and their entire teams. We would also like to acknowledge the contributions of the members of the UPMC Magee Monoclonal Antibody Treatment Group: Rich Beigi, Maribeth McLaughlin, Hyagriv Simhan, Harold Wiesenfeld, Scarlet Lau, Michael Haley, Sandy Trizzino, Ashley Steiner, Lauren Wiser, Michelle Adam, Tina Borneman, David T. Huang, Richard J. Wadas, Russell Meyers, J. Ryan Bariola, Mark Schmidhofer, Graham Snyder, Donald M. Yealy, Derek C. Angus, Tami Minnier, Judith A. Shovel, Debbie Albin, Oscar C. Marroquin, Kevin Collins, Adam King, Kevin E. Kip, Mary Kay Wisniewski, Colleen Sullivan, Meredith Axe, William Garrard, Stephanie Montgomery, Ghady Haidar, Paula L. Kip, Rachel L. Zapf, Sharen Ziska, Jessica Shirley, and Rebecca Medva.

## Author contributions

Dr. McCreary takes full responsibility for the integrity of the data and the accuracy of the data analysis.

*Study concept and design:* All authors

*Acquisition of data:* Lara Lemon, Christina Megli

*Interpretation of data:* All authors

*Drafting of the manuscript:* All authors

*Critical revision of the manuscript for important intellectual content:* All authors

*Study supervision:* All authors

## Summary of Supplements

### Supplement 1 – Supplemental Figures and Tables

**eFigure 1**. Forest Plot of Adjusted Risk Ratios for COVID-19 Associated Outcome from Propensity Score Adjusted Analysis, by Subgroups

**eFigure 2**. Forest Plot of Adjusted Risk Ratios for Non-COVID-19 Associated Outcome from Propensity Score Adjusted Analysis, by Subgroups

**eFigure 3**. Cumulative Density Plot of Non-COVID-19 Related Outcome Events, Stratified by Monoclonal Antibody Treatment

**eFigure 4**. Histograms of Propensity Scores, Stratified by Monoclonal Antibody Treatment

**eFigure 5**. Love Plot of Standardized Percent Bias Across Covariates Before and After Propensity Score Matching

**eFigure 6**. Histograms of Cohort Accrual Over Time, Stratified by Monoclonal Antibody Treatment

**eFigure 7**. Histograms of Gestational Age at Index Date, Stratified by Monoclonal Antibody Treatment

**eTable 1**. Comparison of Characteristics of Pregnant mAb Treated vs Non-Treated Persons in Propensity Score-Matched Cohorts

**eTable 2**. Risk-Adjusted Frequency of COVID-19 Associated Outcome or Other Admission Within 28-days of SARS-CoV-2 positive Test or Monoclonal Antibody Infusion in Unvaccinated, Vaccinated, Obese and Non-Obese Pregnancies

**eTable 3**. Hypertensive Disorder at Delivery Identified by ICD Coding Documented During Delivery Admission

### Supplement 2 – STROBE and Extended RECORD Checklist

## FIGURE LEGENDS

**Figure 1.**
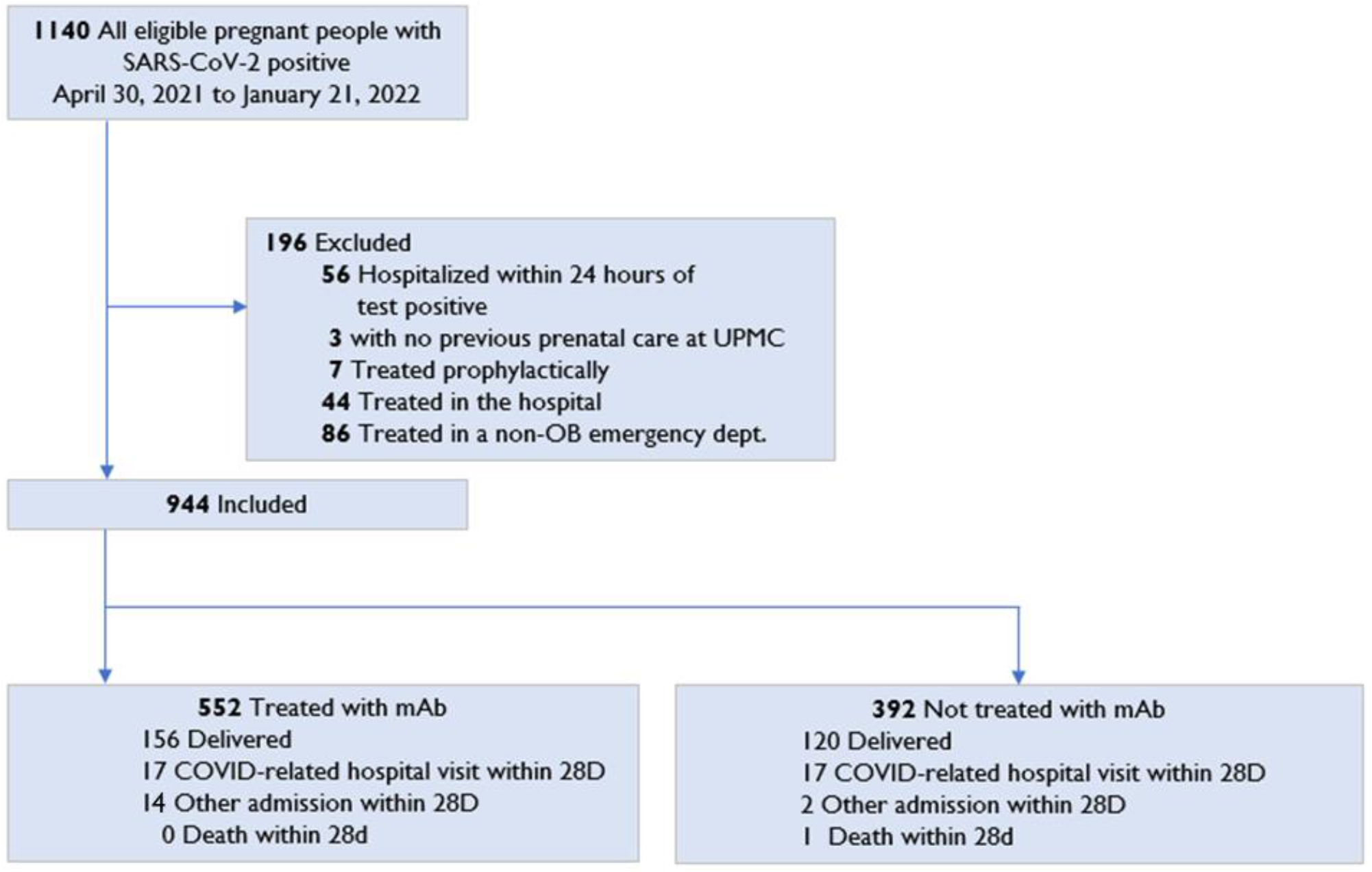
Cohort accrual. Of 1,140 eligible pregnant persons with a SARS-CoV-2 positive test, 944 were mAb eligible, and of these, 552 were treated with mAb and 392 were not treated.

