## Supplement 2 for "Monoclonal Antibodies for Treatment of SARS-CoV-2 Infection During Pregnancy"

### Supplemental STROBE and Extended RECORD Checklist

| Section | Items | STROBE | Page/s | RECORD | Page/s |
|---|---|---|---|---|---|
| **Title and abstract** | | | | | |
| | 1 | (a) Indicate the study's design with a commonly used term in the title or the abstract. (b) Provide in the abstract an informative and balanced summary of what was done and what was found. | | RECORD 1.1: The type of data used should be specified in the title or abstract. When possible, the name of the databases used should be included. RECORD 1.2: If applicable, the geographic region and timeframe within which the study took place should be reported in the title or abstract. | 1-4 |
| **Introduction** | | | | | |
| Background rationale | 2 | Explain the scientific background and rationale for the investigation being reported. | | | 6 |
| Objectives | 3 | State specific objectives, including any prespecified hypotheses. | | | 6 |
| **Methods** | | | | | |
| Study Design | 4 | Present key elements of study design early in the paper. | | | 7-8 |
| Setting | 5 | Describe the setting, locations, and relevant dates, including periods of recruitment, exposure, follow-up, and data collection. | | | 7-8 |

| | | | | |
|---|---|---|---|---|
| Participants | 6 | *Cohort study* - Give the eligibility criteria, and the sources and methods of selection of participants. Describe methods of follow-up | RECORD 6.1: The methods of study population selection (such as codes or algorithms used to identify subjects) should be listed in detail. If this is not possible, an explanation should be provided.<br>RECORD 6.2: Any validation studies of the codes or algorithms used to select the population should be referenced. If validation was conducted for this study and not published elsewhere, detailed methods and results should be provided. | 8 |
| Variables | 7 | Clearly define all outcomes, exposures, predictors, potential confounders, and effect modifiers. Give diagnostic criteria, if applicable. | RECORD 7.1: A complete list of codes and algorithms used to classify exposures, outcomes, confounders, and effect modifiers should be provided. If these cannot be reported, an explanation should be provided. | 8 |
| Data sources/ measurement | 8 | For each variable of interest, give sources of data and details of methods of assessment (measurement). Describe comparability of assessment methods if there is more than one group. | | 9-10 |
| Bias | 9 | Describe any efforts to address potential sources of bias. | | 9-10 |
| Study size | 10 | Explain how the study size was arrived at. | | 9-10 |

| | | | | |
|---|---|---|---|---|
| Quantitative variables | 11 | Explain how quantitative variables were handled in the analyses. If applicable, describe which groupings were chosen, and why. | | 9-10 |
| Statistical methods | 12 | (a) Describe all statistical methods, including those used to control for confounding.<br>(b) Describe any methods used to examine subgroups and interactions.<br>(c) Explain how missing data were addressed. | | 10-11 |
| Data access and cleaning methods | | | RECORD 12.1: Authors should describe the extent to which the investigators had access to the database population used to create the study population.<br>RECORD 12.2: Authors should provide information on the data cleaning methods used in the study. | 10-11 |

| | | | | |
|---|---|---|---|---|
| **Results** | | | | |
| Participants | 13 | (a) Report the numbers of individuals at each stage of the study (*e.g.*, numbers potentially eligible, examined for eligibility, confirmed eligible, included in the study, completing follow-up, and analyzed).<br>(b) Give reasons for non-participation at each stage.<br>(c) Consider use of a flow diagram. | RECORD 13.1: Describe in detail the selection of the persons included in the study (*i.e.,* study population selection) including filtering based on data quality, data availability and linkage. The selection of included persons can be described in the text and/or by means of the study flow diagram. | 12 |
| Descriptive data | 14 | (a) Give characteristics of study participants (*e.g.*, demographic, clinical, social) and information on exposures and potential confounders.<br>(b) Indicate the number of participants with missing data for each variable of interest. | | 12-13 |
| Outcome data | 15 | *Cohort study* - Report numbers of outcome events or summary measures over time. | | 13-14 |

| | | | | |
|---|---|---|---|---|
| Main results | 16 | (a) Give unadjusted estimates and, if applicable, confounder-adjusted estimates and their precision (e.g., 95% confidence interval). Make clear which confounders were adjusted for and why they were included.<br>(b) Report category boundaries when continuous variables were categorized.<br>(c) If relevant, consider translating estimates of relative risk into absolute risk for a meaningful time period. | | 12-14 |
| Other analyses | 17 | Report other analyses done—e.g., analyses of subgroups and interactions, and sensitivity analyses. | | 13-14 |
| **Discussion** | | | | |
| Key results | 18 | Summarize key results with reference to study objectives. | | 15 |
| Limitations | 19 | Discuss limitations of the study, taking into account sources of potential bias or imprecision. Discuss both direction and magnitude of any potential bias. | RECORD 19.1: Discuss the implications of using data that were not created or collected to answer the specific research question(s). Include discussion of misclassification bias, unmeasured confounding, missing data, and changing eligibility over time, as they pertain to the study being reported. | 17 |
| Interpretation | 20 | Give a cautious overall interpretation of results considering objectives, limitations, multiplicity of analyses, results from similar studies, and other relevant evidence. | | 15-16 |

| | | | | |
|---|---|---|---|---|
| Generalizability | 21 | Discuss the generalizability (external validity) of the study results. | | 16 |
| **Other Information** | | | | |
| Funding | 22 | Give the source of funding and the role of the funders for the present study and, if applicable, for the original study on which the present article is based. | | 20 |
| Accessibility of protocol, raw data, and programming code | | .. | RECORD 22.1: Authors should provide information on how to access any supplemental information such as the study protocol, raw data, or programming code. | N/A |
