## Supplement 1 for "Monoclonal Antibodies for Treatment of SARS-CoV-2 Infection During Pregnancy"

### Table of Contents


| | | |
|---|---|---|
| **eFigure 1**. | Forest Plot of Adjusted Risk Ratios for COVID-19 Associated Outcome from Propensity Score Adjusted Analysis, by Subgroups | **2** |
| **eFigure 2**. | Forest Plot of Adjusted Risk Ratios for Non-COVID-19 Associated Outcome from Propensity Score Adjusted Analysis, by Subgroups | **3** |
| **eFigure 3**. | Cumulative Density Plot of Non-COVID-19 Related Outcome Events, Stratified by Monoclonal Antibody Treatment | **4** |
| **eFigure 4**. | Histograms of Propensity Scores, Stratified by Monoclonal Antibody Treatment | **5** |
| **eFigure 5**. | Love Plot of Standardized Percent Bias Across Covariates Before and After Propensity Score Matching | **6** |
| **eFigure 6**. | Histograms of Cohort Accrual Over Time, Stratified by Monoclonal Antibody Treatment | **7** |
| **eFigure 7**. | Histograms of Gestational Age at Index Date, Stratified by Monoclonal Antibody Treatment | **8** |
| **eTable 1.** | Comparison of Characteristics of Pregnant mAb Treated vs Non-Treated Persons in Propensity Score-Matched Cohorts | **9** |
| **eTable 2.** | Risk-Adjusted Frequency of COVID-19 Associated Outcome or Other Admission Within 28-days of SARS-CoV-2 positive Test or Monoclonal Antibody Infusion in Unvaccinated, Vaccinated, Obese and Non-Obese Pregnancies | **10** |
| **eTable 3.** | Hypertensive Disorder at Delivery Identified by ICD Coding Documented During Delivery Admission | **11** |

**eFigure 1. Forest Plot of Adjusted Risk Ratios for COVID-19 Associated Outcome from Propensity Score Adjusted Analysis, by Subgroups**

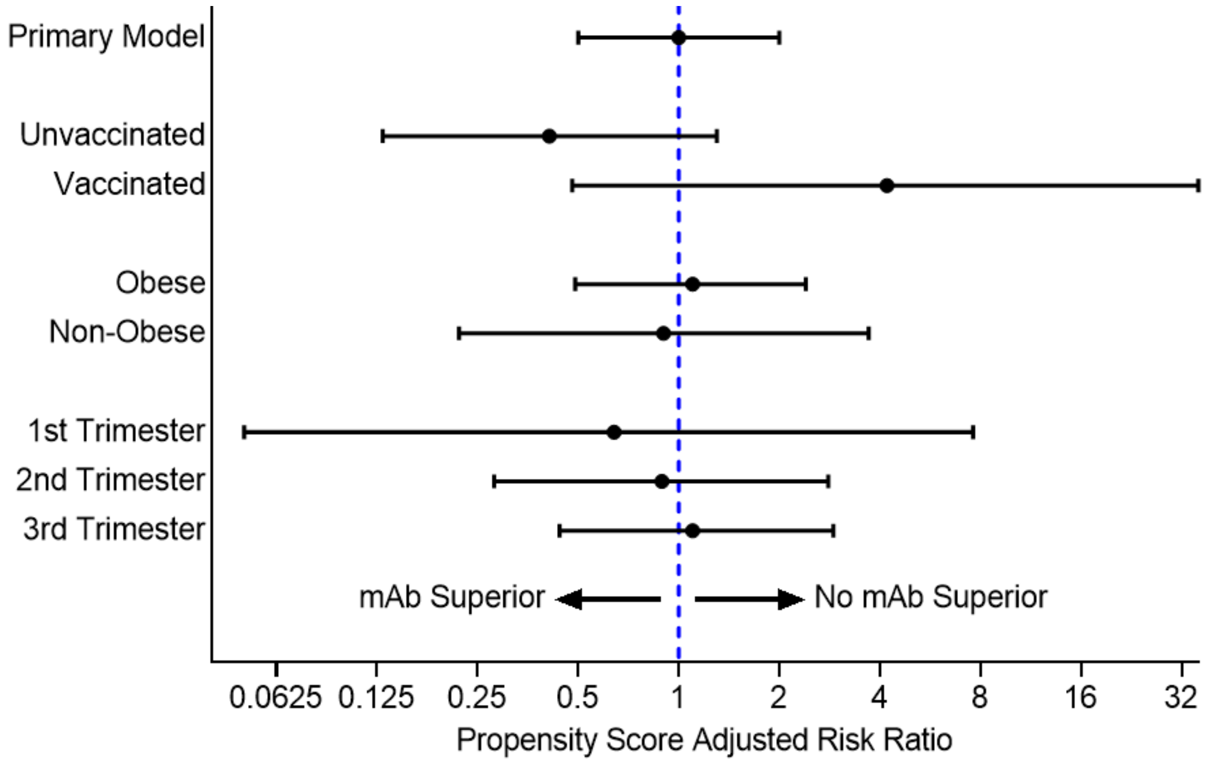


First trimester= <14 weeks gestation
Second trimester=14 weeks to <27 weeks gestation
Third trimester= 27+ weeks gestation

*Abbreviations*: mAb: monoclonal antibodies

**eFigure 2. Forest Plot of Adjusted Risk Ratios for Non-COVID-19 Associated Outcome from Propensity Score Adjusted Analysis, by Subgroups**

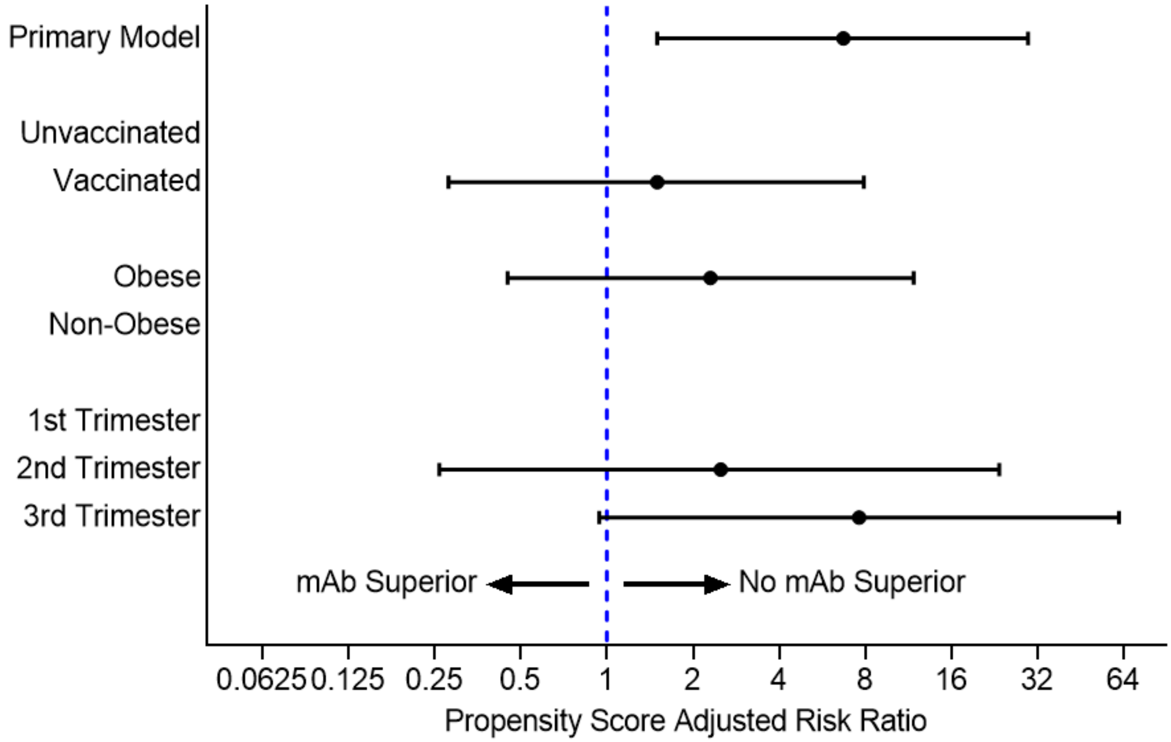


First trimester= <14 weeks gestation
Second trimester=14 weeks to <27 weeks gestation
Third trimester= 27+ weeks gestation

*Abbreviations*: mAb: monoclonal antibodies

**eFigure 3. Cumulative Density Plot of Non-COVID-19 Related Outcome Events, Stratified by Monoclonal Antibody Treatment**

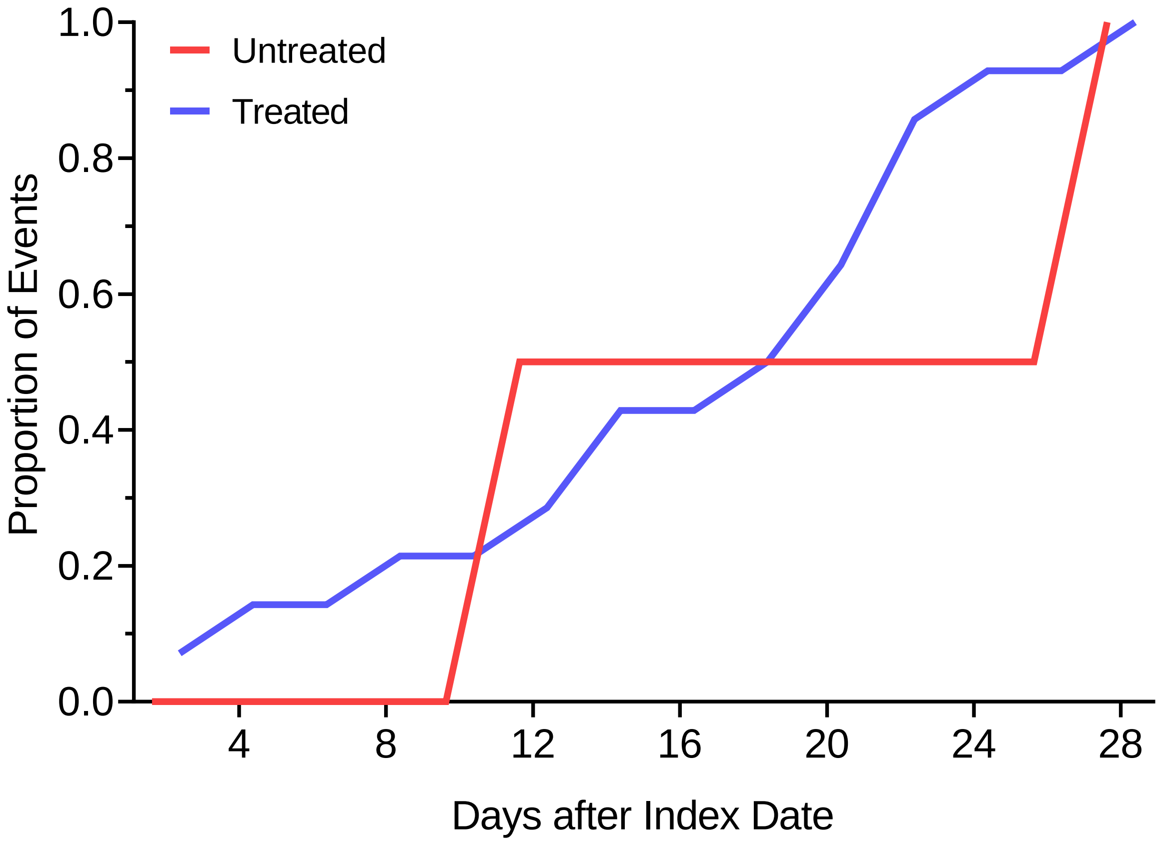


Red represents untreated, blue represents treated with monoclonal antibodies.

### eFigure 4. Histograms of Propensity Scores, Stratified by Monoclonal Antibody Treatment

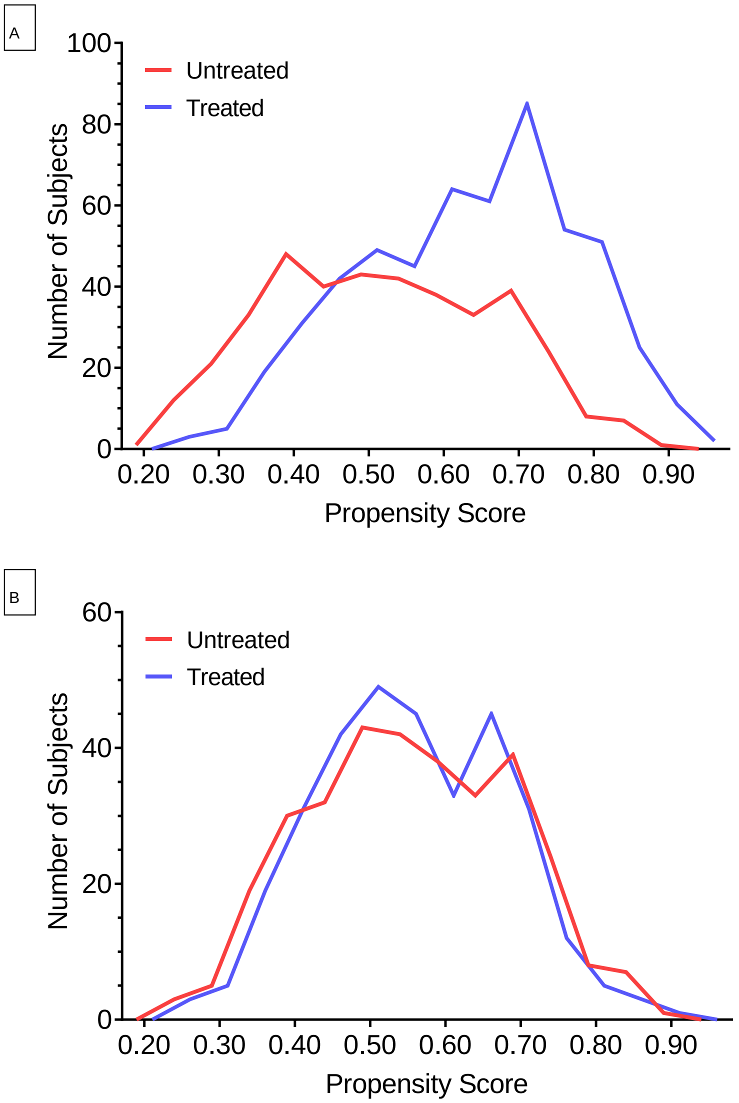


Red represents untreated, blue represents treated with monoclonal antibodies. Panel A represents a histogram of propensity scores for the full cohort (n=944). Panel B represents a histogram of propensity scores for the matched cohort (n=648).

**eFigure 5. Love Plot of Standardized Percent Bias Across Covariates Before and After Propensity Score Matching**

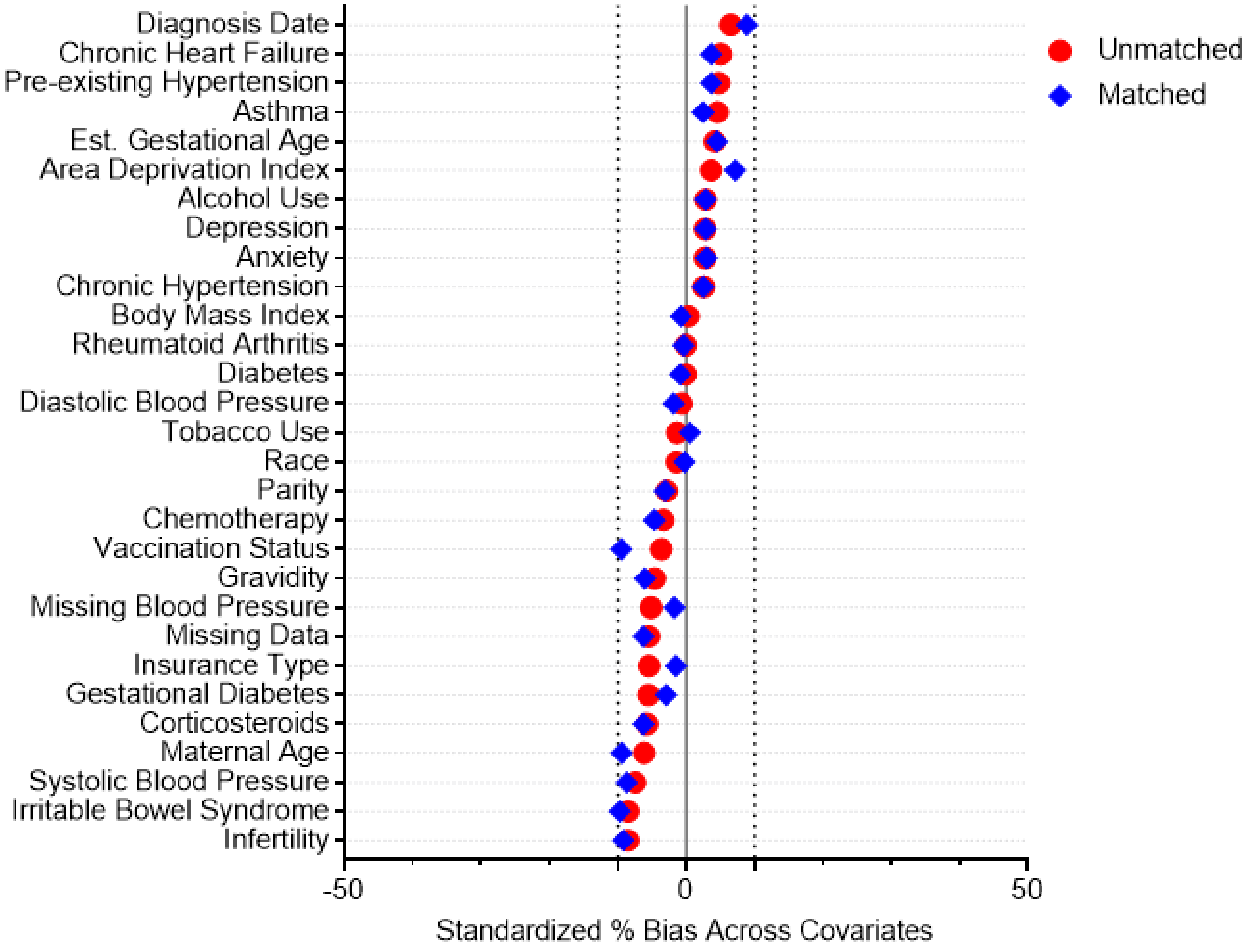


Red circle represents % bias across covariates before propensity score matching. Blue diamond represents % bias across covariates after propensity score matching.

**eFigure 6. Histograms of Cohort Accrual Over Time, Stratified by Monoclonal Antibody Treatment**

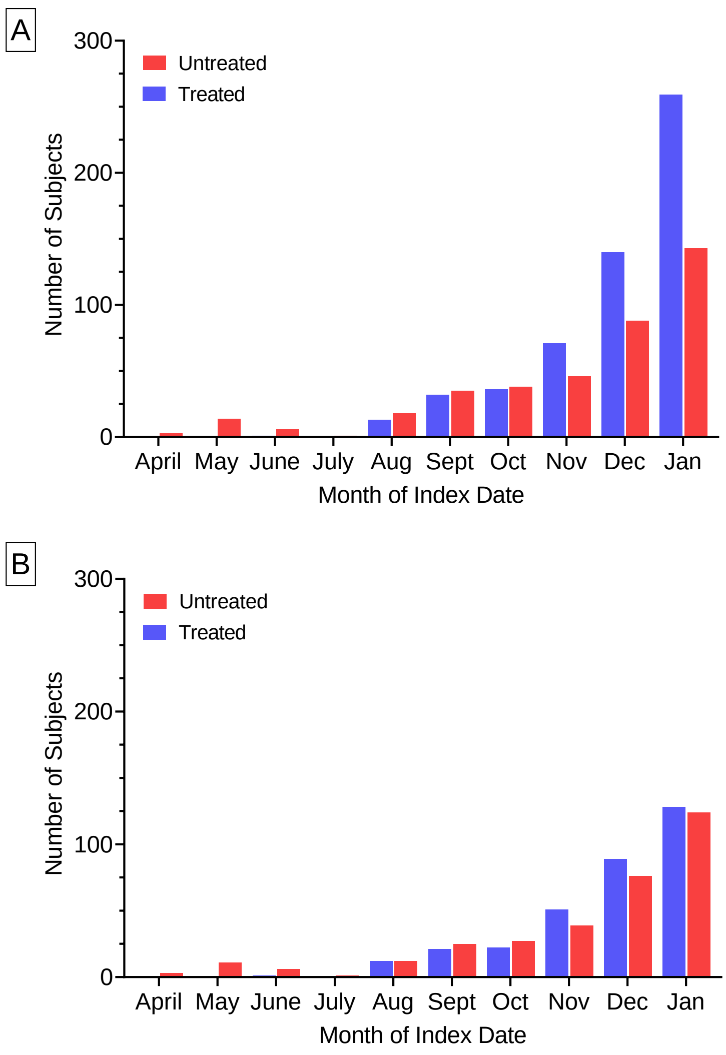


Red represents untreated and blue represents treated with monoclonal antibodies. Panel A represents a histogram of accrual of the full cohort (n=944). Panel B represents a histogram of accrual for the propensity matched cohort (n=648).

### eFigure 7. Histograms of Gestational Age at Index Date, Stratified by Monoclonal Antibody Treatment

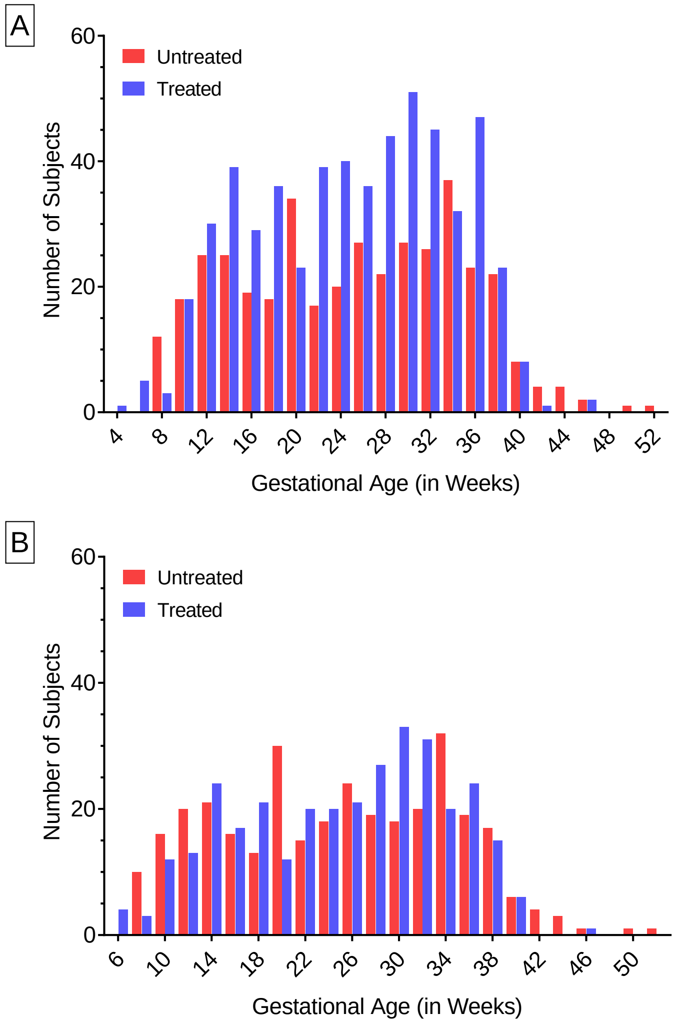


Red represents untreated and blue represents treated with monoclonal antibodies. Panel A represents a histogram of gestational age at index date for the full cohort (n=944). Panel B represents a histogram of gestational age at index date for the propensity matched cohort (n=648).

**eTable 1. Comparison of Characteristics of Pregnant mAb Treated vs. Non-treated Persons in Propensity Score-Matched Cohort**

| | All Patients | Treated | Non-Treated | p-value |
|---|---|---|---|---|
| No. | 648 | 324 | 324 | |
| *Patient Characteristics* | | | | |
| Age, Median (IQR) years | 29.1 (25.2, 33.0) | 29.0 (24.0, 33.0) | 29.4 (25.8, 33.0) | 0.37 |
| Race, n (%) | | | | |
| White | 494 (76.2%) | 246 (75.9%) | 248 (76.5%) | 0.90 |
| Black | 134 (20.7%) | 67 (20.7%) | 67 (20.7%) | |
| Other | 20 (3.1%) | 11 (3.4%) | 9 (2.8%) | |
| Medical History, n (%) | | | | |
| Smoking | 210 (32.4%) | 104 (32.1%) | 106 (32.7%) | 0.87 |
| Diabetes-Pre-gestational | 10 (1.5%) | 5 (1.5%) | 5 (1.5%) | 1.00 |
| Diabetes-Gestational | 19 (2.9%) | 8 (2.5%) | 11 (3.4%) | 0.48 |
| Asthma | 211 (32.6%) | 109 (33.6%) | 102 (31.5%) | 0.56 |
| Hypertension-Preexisting | 25 (3.9%) | 14 (4.3%) | 11 (3.4%) | 0.54 |
| Hypertension-Preeclampsia | 0 (0) | 0 (0) | 0 (0.0) | NA |
| Infertility | 45 (6.9%) | 19 (5.9%) | 26 (8.0%) | 0.28 |
| Other obstetric conditions | 40 (6.2%) | 22 (6.8%) | 18 (5.6%) | 0.51 |
| Cancer | 2 (03%) | 0 (0.0%) | 2 (0.6%) | 0.16 |
| Chemotherapy | 24 (3.7%) | 11 (3.4%) | 13 (4.0%) | 0.68 |
| Other chronic conditions | 38 (5.9%) | 20 (6.2%) | 18 (5.6%) | 0.74 |
| Corticosteroid treatment | 76 (11.7%) | 35 (10.8%) | 41 (12.7%) | 0.46 |
| Vaccination status, n (%) | | | | |
| Unvaccinated | 143 (22.1%) | 91 (28.1%) | 52 (16.0%) | 0.001 |
| Partially vaccinated | 40 (6.2%) | 23 (7.1%) | 17 (5.2%) | |
| Fully vaccinated | 222 (34.3%) | 100 (30.9%) | 122 (37.7%) | |
| Unknown/not determined | 243 (37.5%) | 110 (34.0%) | 133 (41.0%) | |
| Charlson Comorbidity Index Score, Median (IQR) | 0 (0, 0) | 0 (0, 0) | 0 (0, 0) | 0.94 |
| Body mass index, Median (IQR) | 30.2 (26.6, 34.3) | 31 (26.7, 33.6) | 30 (26.4, 34.5) | 0.77 |
| Gestational age at COVID-19 diagnosis or infusion (days), Median (IQR) | 180.5 (120.0, 228.5) | 187 (125, 223.5) | 175.5 (117.5, 231.5) | 0.51 |
| Trimester, n (%) | | | | |
| First trimester | 102 (15.7%) | 43 (13.3%) | 59 (18.2%) | 0.19 |
| Second trimester | 248 (38.3%) | 124 (38.3%) | 124 (38.3%) | |
| Third trimester | 298 (46.0%) | 157 (48.5%) | 141 (43.5%) | |
| Nulliparous, n (%) | 223 (34.4%) | 113 (34.9%) | 110 (34.0%) | 0.80 |
| Early miscarriage, n (%) | 6 (0.9%) | 4 (1.2%) | 2 (0.6%) | 0.41 |
| Insurance type, n (%) | | | | |
| Commercial | 381 (58.8%) | 193 (59.6%) | 188 (58.0%) | 0.65 |
| Public | 221 (34.1%) | 111 (34.3%) | 110 (34.0%) | |
| Self-pay/Other | 46 (7.1%) | 20 (6.2%) | 26 (8.0%) | |
| Area Deprivation Index, Median (IQR) | 71 (53, 86) | 70 (53, 86) | 71 (53, 86) | 0.77 |

**eTable 2. Risk-Adjusted Frequency of COVID-19 Associated Outcome or Other Admission Within 28-days of SARS-CoV-2 Positive Test or Monoclonal Antibody Infusion in Unvaccinated, Vaccinated, Obese, and Non-Obese Pregnancies**

| Model | No. | Total Visits/Total No. (%) | | Risk-Adjusted 28-day COVID visit, per 100 people (95%CI) | | Risk Difference, per 100 people (95% CI) | Risk Ratio (95% CI) |
|---|---|---|---|---|---|---|---|
| | | Treated | Non-treated | Treated | Non-treated | | |
| *Unvaccinated* | | | | | | | |
| COVID-associated | | | | | | | |
| Crude | 178 | 5/122 (4.1%) | 6/56 (10.7%) | 4.1 (0.58, 7.6) | 10.7 (2.6, 18.8) | -6.6 (-15.5, 2.2) | 0.38 (0.12, 1.2) |
| PS-adjusted | 177 | 5/121 (4.1%) | 6/56 (10.7%) | 4.2 (0.61, 7.8) | 10.2 (2.5, 18.0) | -6.0 (-14.5, 2.5) | 0.41 (0.13, 1.3) |
| Other admission | | | | | | | |
| Crude | 178 | 6/122 (4.9%) | 0 | --- | --- | --- | --- |
| PS-adjusted | 177 | 6/121 (5.0%) | 0 | --- | --- | --- | --- |
| *Vaccinated* | | | | | | | |
| COVID-associated | | | | | | | |
| Crude | 392 | 6/265 (2.3%) | 1/127 (0.79%) | 2.3 (0.47, 4.1) | 0.79 (-0.75, 2.3) | 1.5 (-0.88, 3.8) | 2.9 (0.35, 23.6) |
| PS-adjusted | 387 | 6/262 (2.3%) | 1/125 (0.8%) | 2.6 (0.53, 4.7) | 0.63 (-0.61, 1.9) | 2.0 (-0.47, 4.4) | 4.2 (0.48, 35.9) |
| Other admission | | | | | | | |
| Crude | 392 | 6/265 (2.3%) | 2/127 (1.6%) | 2.3 (0.47, 4.1) | 1.6 (-0.59, 3.7) | 0.69 (-2.1, 3.5) | 1.4 (0.29, 7.0) |
| PS-adjusted | 387 | 6/262 (2.3%) | 2/125 (1.6%) | 2.3 (0.47, 4.2) | 1.5 (-0.61, 3.7) | 0.80 (-2.1, 3.7) | 1.5 (0.30, 7.7) |
| *Obese* | | | | | | | |
| COVID-associated | | | | | | | |
| Crude | 569 | 13/337 (3.9%) | 13/232 (5.6%) | 3.9 (1.8, 5.9) | 5.6 (2.6, 8.6) | -1.8 (-5.4, 1.9) | 0.69 (0.33, 1.5) |
| PS-adjusted | 560 | 13/332 (3.9%) | 12/228 (5.3%) | 4.7 (2.2, 7.2) | 4.3 (1.8, 6.7) | 0.39 (-3.2, 4.0) | 1.1 (0.49, 2.4) |
| Other admission | | | | | | | |
| Crude | 569 | 10/337 (3.0%) | 2/232 (0.86%) | 3.0 (1.2, 4.8) | 0.86 (-0.33, 2.1) | 2.1 (-0.06, 4.3) | 3.4 (0.76, 15.6) |
| PS-adjusted | 560 | 10/332 (3.0%) | 2/228 (0.88%) | 3.0 (1.1, 4.9) | 0.88 (-0.35, 2.1) | 2.1 (-0.19, 4.4) | 3.4 (0.71, 16.1) |
| *Non-Obese* | | | | | | | |
| COVID-associated | | | | | | | |
| Crude | 375 | 4/215 (1.9%) | 4/160 (2.5%) | 1.9 (0.05, 3.7) | 2.5 (0.08, 4.9) | -0.6 (-3.7, 2.4) | 0.74 (0.19, 2.9) |
| PS-adjusted | 370 | 4/215 (1.9%) | 4/155 (2.6%) | 2.1 (0.04, 4.1) | 2.3 (0.03, 4.5) | -0.22 (-3.3, 2.8) | 0.90 (0.22, 3.7) |
| Other admission | | | | | | | |
| Crude | 375 | 7/215 (3.3%) | 0 | --- | --- | --- | --- |
| PS-adjusted | 370 | 7/215 (3.3%) | 0 | --- | --- | --- | --- |

**eTable 3. Hypertensive Disorder At Delivery Identified by the following ICD Coding Documented During Delivery Admission**

| Codes: | | | | | | | | | |
|---|---|---|---|---|---|---|---|---|---|
| O10011 | O10919 | O1492 | O1493 | O1494 | O1495 | O1500 | O1502 | O1503 | O151 |
| O10012 | O1092 | O152 | O159 | O161 | O162 | O163 | O164 | O165 | O169 |
| O10013 | O1093 | O1422 | O1042 | O1412 | O1032 | O1402 | O1022 | O133 | O10119 |
| O10019 | O111 | O1423 | O1043 | O1413 | O1033 | O1403 | O1023 | O134 | O10211 |
| O1002 | O112 | O1424 | O10911 | O1414 | O10411 | O1404 | O10311 | O135 | O10212 |
| O1003 | O113 | O1425 | O10912 | O1415 | O10412 | O1405 | O10312 | O139 | O10213 |
| O10111 | O114 | O1490 | O10913 | O1420 | O10413 | O1410 | O10313 | O1400 | |
| O10112 | O115 | O10113 | O119 | O1012 | O131 | O1013 | O132 | O10219 | |
